# Reactive Risk Communication and Media Framing During Nigeria’s 2024 Cholera Outbreak

**DOI:** 10.64898/2026.03.02.26347445

**Authors:** Onuoya J. Ikiba

## Abstract

**Background:** Risk communication is critical in shaping public response during infectious disease outbreaks. This study quantitatively examined whether Nigerian media coverage during the 2024 cholera outbreak reflected a proactive or reactive risk communication pattern.

**Methods:** A Python-based systematic content analysis was conducted on 352 unique news articles published by major Nigerian media sources in 2024. K-Means was used to cluster and quantify thematic patterns, TextBlob for sentiment polarity, and time-series analysis to determine the features of media engagement.

**Results:** The analysis identified a dominant reactive, crisis-driven communication pattern with media coverage surging by over 400% in June, matching the peak of reported cholera cases. Thematic analysis portrayed a severe reporting imbalance focused on Outbreak Reports and Mortality (41.5% of articles), while structural and preventive themes such as WASH and Health Education received marginal attention (less than 25% of coverage). Furthermore, communication was overwhelmingly neutral (76.4%) in sentiment, potentially limiting the perceived urgency required for public action.

**Conclusions:** Media reporting on the 2024 cholera outbreak in Nigeria was reactive and focused disproportionately on threat rather than solutions. These findings support the need for a strategic dual-focus communication model that shifts from crisis-driven coverage to sustained, year-round preventive messaging centered on WASH accountability and community resilience.

## 1. Introduction

Cholera, an acute diarrhoeal illness caused by the Gram-negative bacterium, *Vibrio cholerae,* has evolved from a historical plague in the early 1800s to a determinant of global health inequity [1]. It is a major public health concern in most parts of the developing world, particularly Africa and Asia [2]. In 2017, the Global Task Force on Cholera Control (GTFCC), a global partnership with more than 50 institutions including governments, academia, non-governmental organizations and UN agencies, launched a global roadmap aimed at reducing cholera deaths by 90% and eliminating the disease in twenty countries by 2030 [3]. However, its persistence as a neglected tropical disease can be linked to longstanding systemic failures in public health policy. [4] notes that behavioural factors such as poor environmental sanitation and personal hygiene, and unhealthy environmental practices such as indiscriminate waste and sewage disposal perpetuates cholera, leading to sporadic outbreaks mostly across African states. In Nigeria, cholera has persisted as an endemic disease since the 1970s, with major outbreaks occurring regularly during the rainy season. One of the most severe outbreaks in terms of deaths occurred in 1991, with 59,478 cases and 7,654 deaths. This resulted in a case fatality rate (CFR) of 12.9%; higher than the World Health Organization’s threshold of <1% [5]. In 2024, Nigeria recorded one of the highest cholera-linked mortality rates in Africa. Between January 1 to November 24, 2024, 702 deaths had occurred out of 19, 178 cases, with a CFR of 3.7% [6]. The Monthly Epidemiological Report on Cholera from the Nigeria Centre for Disease Control (NCDC), covering the period of epidemiological weeks 18 to 22 (29 April – 2 June 2024), showed that 882 suspected cases including 16 deaths had been reported in 30 states. Cumulatively, Bayelsa reported 442 cases accounting for 50% of all suspected cases nationwide [7]. The distribution of cholera burden across the most affected states as at week 22 of 2024 is referenced in **Table 1** below:

**Table 1:** Top 9 States by Cumulative Cholera Cases (Epi Week 22, 2024)

| Rank (No.) | State | Cumulative Cases (n) | Percent of Cumulative Cases (%) |
| --- | --- | --- | --- |
| 1 | Bayelsa | 442 | 50% |
| 2 | Zamfara | 64 | 7% |
| 3 | Abia | 51 | 6% |
| 4 | Bauchi | 46 | 5% |
| 5 | Cross River | 42 | 5% |
| 6 | Ebonyi | 38 | 4% |
| 7 | Delta | 34 | 4% |
| 8 | Katsina | 28 | 3% |
| 9 | Imo | 28 | 2% |
| **Total** |  | **773** | **86%** |
| **SOURCE:** NIGERIA CENTRE FOR DISEASE CONTROL, NCDC, 2024. |  |  |  |

While there appears to be a drop in terms of case fatality rate from 12.9% in the 1990’s to 3.7% in 2024, the burden of disease still exceeds acceptable thresholds [8]. The definitive strategy for cholera prevention lies not only in clinical intervention, but in sustained environmental change via improvements in WASH. It has been argued by [9] that while sanitation measures and increased awareness can be relied upon in the control of epidemics, the role of the media in reducing disease prevalence is notable. Furthermore, when there is a disease outbreak such as cholera, there is the urgency to enlighten citizens on the preventive measures. In this regard, the media plays the role of raising awareness that helps to improve health outcomes by persuading people to practice health-seeking behaviours [9–10]. Information is disseminated to the public via programmes and other creative methods to reach diverse audiences [10, 11]. However, the effectiveness of media communication depends on its underlying structure, a process known as framing [12]. There is evidence pointing to the fact that during a crisis, the media employs a ‘crisis frame.’ A posture focused on sensationalism, death tolls and treatment, often at the expense of preventive strategies [12]. While some context on how national newspapers cover health issues have been provided by [12], and Shittu et al. [13] have reported the coverage of cholera outbreak by two selected Nigerian dailies, there is limited systematic, quantitative research offering an empirical synthesis of media narratives using advanced content-based analysis. Therefore, in this study, a systematic, Python-based content analysis was done to quantify the thematic, tonal and temporal characteristics of public health communication regarding cholera and preventive measures during the 2024 outbreak in Nigeria. The intent being to move beyond stories to data-backed insights that can shape future communication strategies towards proactive, year-round cholera prevention and preparedness for other public health emergencies.

## 2. Methodology

This study utilized a systematic Python-based method to quantify media communication on the 2024 cholera outbreak in Nigeria, conducting a framing analysis across thematic, tonal and temporal dimensions.

### 2.1 Data Collection and Scope

Data was collected from January 1 to December 31, 2024, spanning the period of the outbreak. A custom Python-based news mining pipeline (collect_news.py) was built to fetch news articles from major Nigeria media sources via Google News RSS feeds. The search query included broad and specific keywords to ensure comprehensive retrieval, such as: “cholera outbreak”, “sanitation”, “hygiene”, “WASH”, “cholera prevention”, and “Bayelsa cholera 2024”. This programmatic approach was followed to ensure replicability and minimization of selection bias. The initial search retrieved 415 unique news articles.

### 2.2 Data Pre-processing and Analysis Pipeline

The raw data was cleaned through a cleaning pipeline (clean_news.py) using the pandas and Natural Language Toolkit (NLTK) Python libraries. This process removed duplicates, filtered out non-relevant content, and performed text normalization (lowercasing, removal of stop words and stemming). The final article set for analysis comprised 352 usable news articles. The analysis was implemented using Python (v3.13.7) with Scikit-learn, TextBlob and Matplotlib.

### 2.3 Content Analysis and Framing

The core analysis implemented three distinct analytical dimensions (thematic, tonal and temporal) using a multi-level data analysis script (analyze_news.py). To identify news frames, text data was transformed using Term Frequency-Inverse Document Frequency (TF-IDF) vectorization. An unsupervised clustering approach was used to allow thematic patterns to emerge directly from the data rather than relying on predefined categories. K-Means was preferred for its transparent and computationally efficient method for grouping text based on similarity, making it suitable for analyzing a structured corpus of news articles. The optimal number of clusters (K) was determined using the Elbow Method, which examines the rate of decrease in within-cluster sum of squares to identify a point of diminishing returns in model fit [14].The clusters were then manually reviewed and labeled to operationalize the thematic frames. The tone of communication was assessed using sentiment polarity scoring implemented through the TextBlob library, a lexicon-based natural language processing tool that assigns polarity values to text based on weighted word sentiment. Polarity scores ranging from (−1.0 to +1.0) were categorized into three bins: Positive (> 0.1), Negative (<−0.1) and Neutral (−0.1≤ Polarity ≤0.1). Temporal trends were examined by comparing monthly publication frequency of the 352 articles with official cholera case data reported by the Nigeria Centre for Disease Control (NCDC). Official epidemiological case data were extracted from NCDC situation reports and aggregated monthly to enable descriptive comparison with publication frequency. No inferential correlation modelling was performed, as the analysis focused on temporal alignment and surge detection.

## 3. Results

The systematic content analysis was based on the final 352 articles to quantify the media’s framing of the 2024 cholera outbreak in Nigeria across thematic, tonal and temporal dimensions.

### 3.1 Thematic Analysis

The K-Means clustering algorithm identified unique thematic frames. The distribution indicated a severe reporting imbalance, with a focus on Crisis Frame components, and a marginal coverage on Prevention Frame components. The thematic analysis showed a clear disparity in media focus. The Outbreak Reports and Mortality cluster was the dominant theme, accounting for 146 articles (41.5%) of the total sample. The Community Response and Aid theme was next in order with 84 articles (23.9%). In contrast, foundational preventive themes such as Water, Sanitation and Hygiene, received less attention, appearing in only 29 news articles (8.2%). This pattern of reporting is suggestive of the fact that media focus regarding the 2024 cholera outbreak in Nigeria, largely tilted towards immediate crisis updates than long-term, systemic preventive measures. This disproportionate distribution is visualized in **Figure 1**.

**Figure 1:**
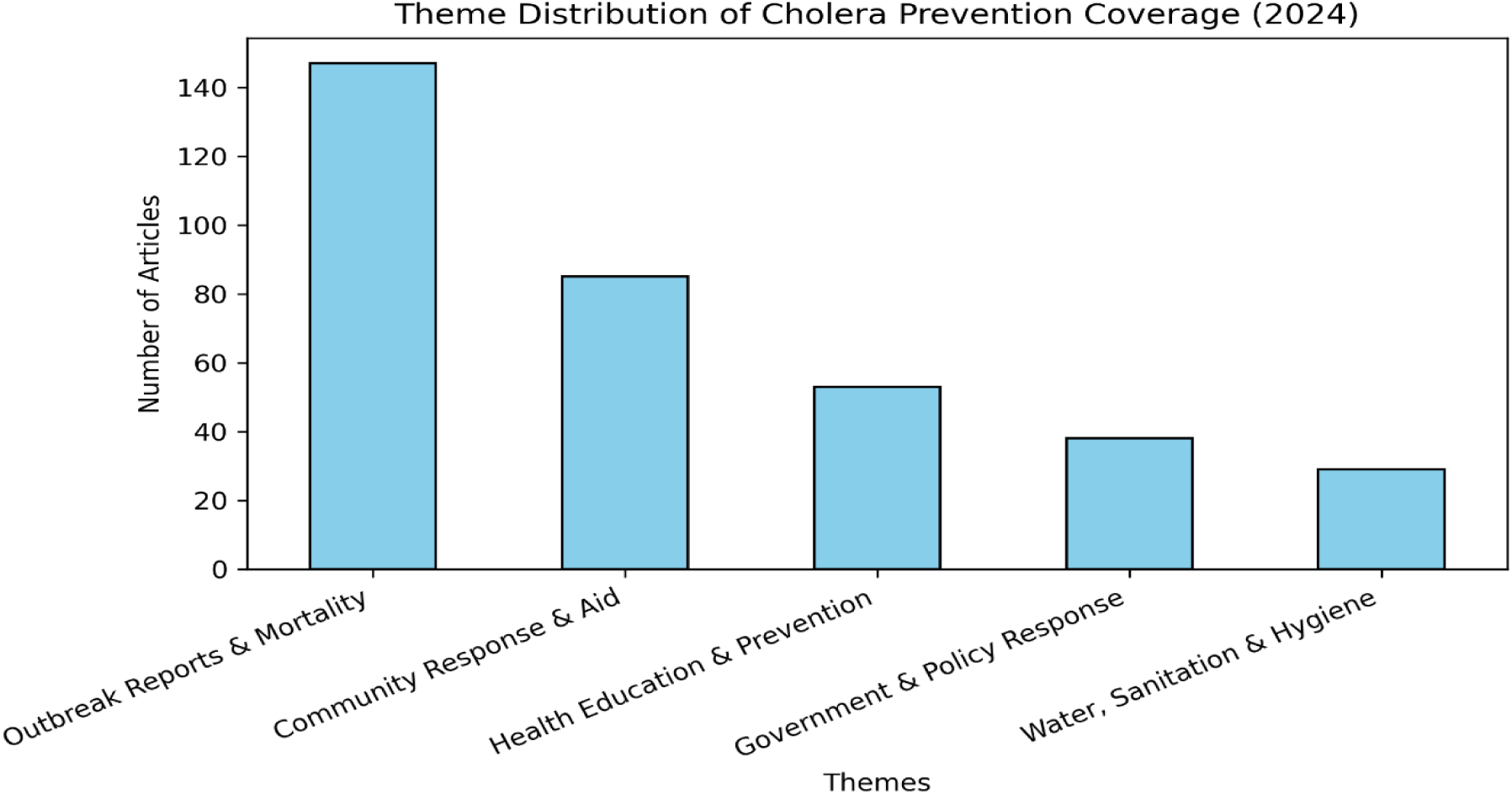
Distribution of cholera-related communication themes in Nigeria (2024)

### 3.2 Tonal Analysis

The tone of publications during the 2024 cholera outbreak in Nigeria was calculated for sentiment polarity using TextBlob library. The pie chart (**Figure 2)** displays the sentiment distribution.

**Figure 2:**
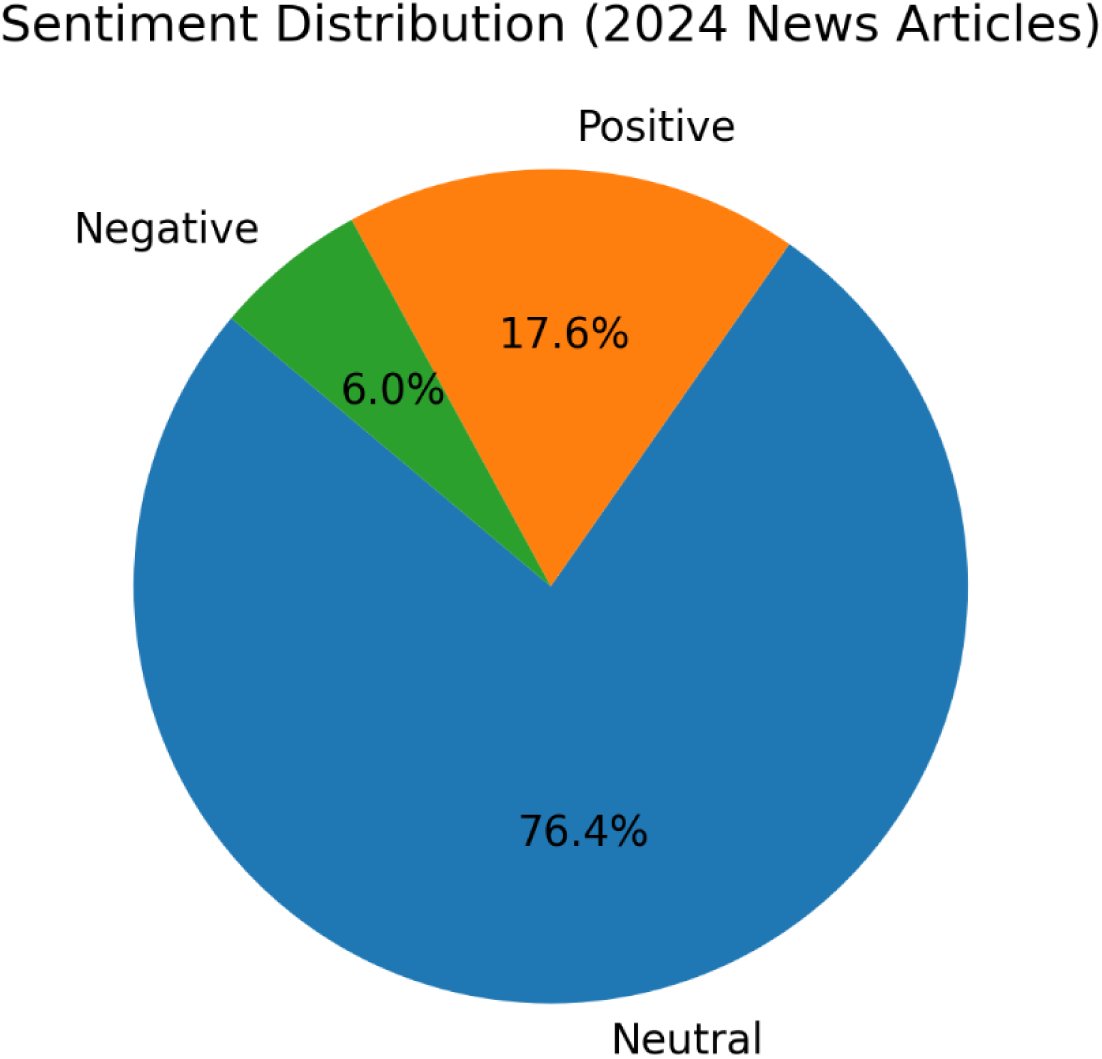
Sentiment distribution of cholera-related news articles in Nigeria (2024)

Sentiment analysis showed that the majority of public health communication adopted a Neutral tone (76.4% of all articles). Articles containing a Positive sentiment comprised 17.6% of the coverage, while those with a Negative sentiment, was a minor 6.0%. This overwhelming neutrality shows that news communication placed more emphasis on factual, descriptive reporting, but may have limited the perceived urgency necessary to motivate rapid public or policy response concerning immediate environmental risks.

### 3.3 Temporal Analysis

Result of the temporal analysis on the 2024 cholera outbreak in Nigeria is presented in Analysis of the publication frequency against the outbreak timeline revealed a distinct, reactive, crisis-driven communication pattern. News coverage on cholera remained consistently low from January to May 2024, with an average of 10-14 articles per month. This trend abruptly changed in June, coinciding with the peak of reported cholera cases, where publication volume increased to approximately 70 news articles (an increase of over 400% compared to the preceding period). Coverage stabilized at a moderate level (30-40 publications per month) between July and September before a minor resurgence in October. This pattern indicates that media engagement primarily serves as a mirror of the immediate epidemiological crisis rather than a sustained, proactive communication tool.

## 4. Discussion

This study systematically analyzed the features of Nigerian news media publications on cholera prevention in 2024, showing patterns that have critical implications for public health risk communication, particularly, in endemic areas. The core findings reveal an overwhelming focus on threat reporting, a pervasive neutral sentiment and a reactive, crisis-driven communication pattern.

First, the thematic analysis presented in **Figure 1** showed a disproportionate bias towards the theme of “outbreak reports and mortality,” while systemic, preventive themes like “health education and prevention,” government and policy response or “water, sanitation and hygiene”, received minimal coverage. This finding strongly agrees with observations by [15,16] that media discourse during health threats often defaults to a crisis framing model. These findings align with crisis framing theory, which suggests that media narratives during emergencies prioritize immediacy and mortality metrics over structural determinants. For instance, reports during the early phases of the COVID-19 pandemic globally concentrated heavily on fear and scaremongering framing, with mentions of fatality rates significantly overshadowing other aspects of disease management [16]. Since media framing of a health crisis can either moderate or escalate its effect on a population, [15] points out that the media must exercise prudence to avoid presenting news in a way that generates unnecessary panic or complicates the overall public health response. In the context of the cholera outbreak in Nigeria, the consequence of this consistent crisis-focus is a detrimental shift in public attention away from underlying systemic issues and essential preventive efforts that are critical to reducing the endemic disease burden. As illustrated in **Table 1**, Bayelsa State’s disproportionate contribution (50% of the cumulative caseload between April and June 2024), warrants the urgent need for prioritized, long-term preventive messaging in endemic areas. Unbalanced or underreporting of infectious disease outbreaks is a recognized challenge in public health research. As [17] notes, this is often influenced by country-level factors, including weak public health infrastructure, issues of government censorship and even inherent media biases concerning perceived severity or stigma. Given that the primary drivers of cholera in Nigeria are intrinsically linked to deficient water, sanitation and hygiene infrastructure [2,5], the media’s minimal coverage of this foundational theme represents a missed opportunity to set the public health agenda, advocate for improved infrastructure, demand accountability and promote sustainable behavioural change, rather than the overwhelming focus on casualty count.

Second, this study critically examined the tone of public health communication in the Nigerian media during the 2024 cholera outbreak. The sentiment analysis result (**Figure 2**) indicates a predominant leaning towards a neutral tone, observed in about three-quarters of the news articles analyzed. While this neutral tone reflects a focus on low-bias, factual reporting, a generally acceptable goal for increasing knowledge and health risk perception during an epidemic [18–19], however, when reporting a health threat, adopting a pervasive neutral tone can inadvertently diminish the perceived urgency of the crisis, thereby weakening the motivational intensity required to produce widespread and sustained behavioural changes towards preventive practices [18,20]. Also, the observed low positive sentiment in the reporting suggests a failure to engender community resilience and self-efficacy. As [21] argues, positive news framing is crucial as it encourages and motivates individuals to adopt protective behaviours during major disease outbreaks, highlighting a gap in the emotional messaging strategy of the media during the 2024 cholera outbreak in Nigeria.

Third and lastly, the temporal analysis of health messaging trends (**Figure 3**) provides compelling evidence that media attention to the 2024 cholera outbreak was overwhelmingly crisis-driven and reactive. Between January and May, coverage was at a baseline low of fewer than 15 news articles. In sharp contrast, the month of June witnessed a dramatic increase in publications, corresponding to the surge in national cholera caseloads reported during that period. This pattern confirms that media engagement was triggered by the escalation of the epidemic rather than being initiated in anticipation of an outbreak or in response to early warning signals. A subsequent decline in coverage intensity between July and September, followed by a slight rise in October, suggests that reporting remained tied to renewed outbreak alerts or official updates. This reactive pattern demonstrates a critical failing in proactive risk communication, limiting the media’s capacity to build and sustain public awareness, preparedness and preventive behaviour during either pre-outbreak periods or the crucial declining phase of the epidemic, a position also supported by [22]. Collectively, the thematic imbalance, dominant neutrality in tone, and sharp temporal surge during peak caseload months indicate that media engagement functioned primarily as a reactive mirror of epidemiological escalation rather than a sustained risk communication instrument. The ability to systematically quantify and link media focus directly to public health priorities during a national outbreak, along with the real-time systematic analysis of communication characteristics during an active epidemiologic event are key methodological strengths of this study. However, limitations include the exclusion of social media, a vital and influential channel for crisis communication, and the exclusive reliance on the output of news articles alone rather than incorporating broader outcomes like behavioural changes. In addition, the reliance on lexicon-based sentiment analysis may underestimate contextual framing nuances, and clustering outputs remain sensitive to preprocessing decisions and cluster parameter selection. Based on these findings, it is strongly recommended that the NCDC assume a more directive, steering position to shift health communication away from a reactive approach to a proactive and dual-focus model. This requires establishing strategic and sustained partnerships between the media and major public health stakeholders. Practical steps should include sustained, year-round media outreaches, adoption of a balanced framing of messaging that combines threat with efficacy, and implementation of high-intensity communication campaigns especially targeting highly endemic regions to drive lasting behavioural change.

**Figure 3:**
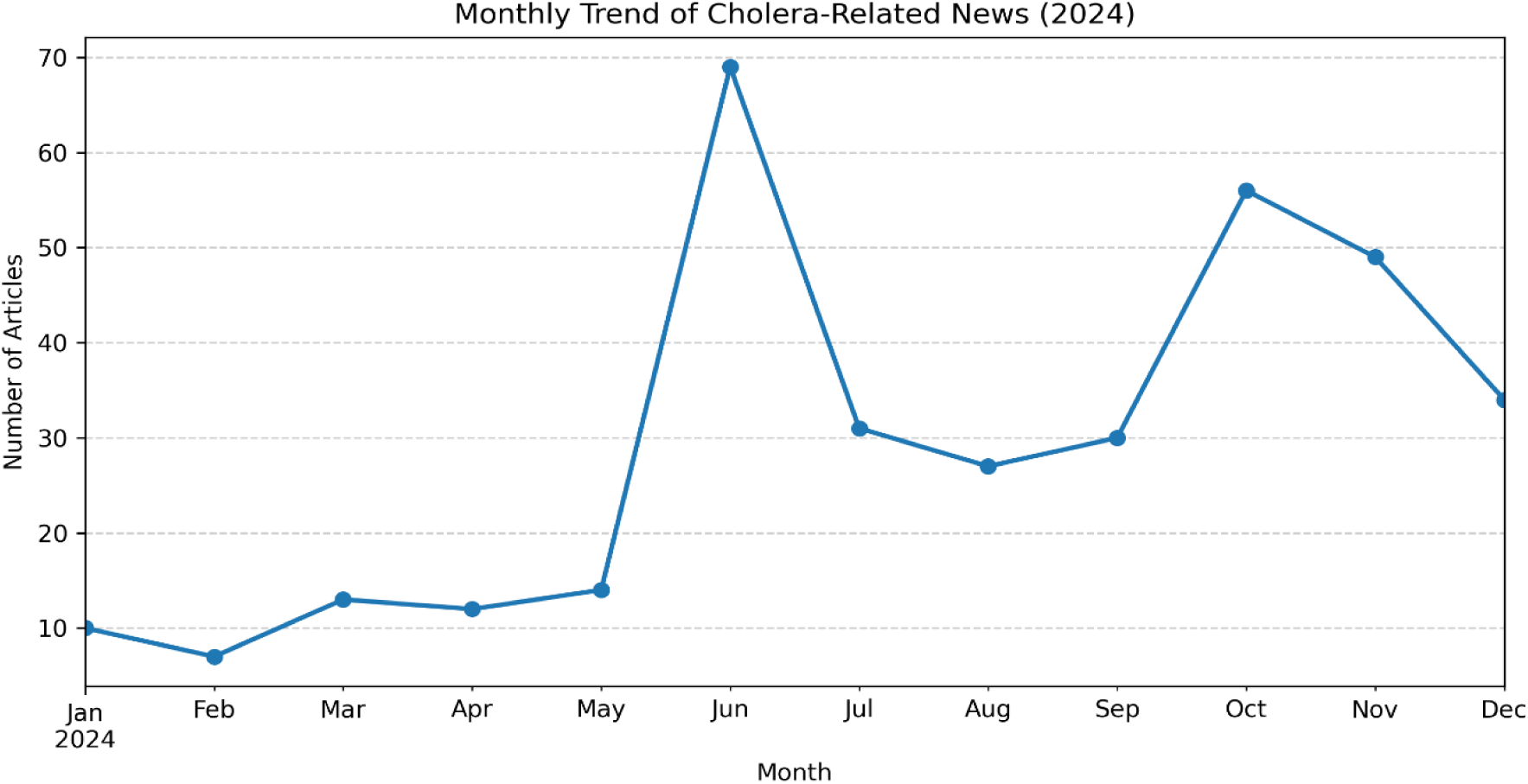
Monthly trend of cholera-related news coverage in Nigeria (2024)

## 5. Conclusions

In conclusion, the systematic content analysis of media coverage during the 2024 cholera outbreak shows that the Nigerian news media’s role was overwhelmingly reactive, focused primarily on crisis reporting (mortality and caseloads) with a pervasive neutral tone. This pattern appears to underemphasize systemic drivers of endemic transmission. To effectively break this persistent cycle of cholera outbreaks, future national public health communication strategies must shift from a reactive to a proactive, sustained model that prioritizes results-oriented messaging that compels the adoption of necessary WASH preventive behaviours.

## Data Availability

The data generated and analyzed during this study are publicly available in the GitHub repository provided in the linked URL https://github.com/Fondestjoel/WASH_Cholera_2024_Outbreak_Project.

## 6 Acknowledgements

Not Applicable

## 7. Disclosure Statements

### Ethics approval and consent to participate

This study used publicly available articles from Google News feeds and did not involve human participants, personal data, or confidential information. As such, ethical approval from an institutional review board was not required.

### Availability of data and materials

The data generated and analyzed during this study are publicly available in the GitHub repository:https://github.com/Fondestjoel/WASH_Cholera_2024_Outbreak_Project.

Researchers can access and reuse these materials in accordance with the repository’s terms. All scripts, preprocessing pipelines, and analysis parameters are documented to enable full methodological replication.

### Authors’ contributions

This work from conception to final output was done solely by this author.

